# Urinary Metal Levels and Coronary Artery Calcification: Longitudinal Evidence in the Multi-Ethnic Study of Atherosclerosis (MESA)

**DOI:** 10.1101/2023.10.31.23297878

**Authors:** Katlyn E. McGraw, Kathrin Schilling, Ronald A. Glabonjat, Marta Galvez-Fernandez, Arce Domingo-Relloso, Irene Martinez-Morata, Miranda R. Jones, Wendy S. Post, Joel Kaufman, Maria Tellez-Plaza, Linda Valeri, Elizabeth R. Brown, Richard A. Kronmal, Graham R. Barr, Steven Shea, Ana Navas-Acien, Tiffany R. Sanchez

**Author notes:** Correspondence: Katlyn E McGraw, PhD, Columbia University Mailman School of Public Health, Department of Environmental Health Science, 722 W 168th St, New York, NY 10032.

## Abstract

**Objective:** Growing evidence indicates that exposure to metals are risk factors for cardiovascular disease (CVD). We hypothesized that higher urinary levels of metals with prior evidence of an association with CVD, including non-essential (cadmium, tungsten, and uranium) and essential (cobalt, copper, and zinc) metals are associated with baseline and rate of change of coronary artery calcium (CAC) progression, a subclinical marker of atherosclerotic CVD.

**Methods:** We analyzed data from 6,418 participants in the Multi-Ethnic Study of Atherosclerosis (MESA) with spot urinary metal levels at baseline (2000-2002) and 1-4 repeated measures of spatially weighted coronary calcium score (SWCS) over a ten-year period. SWCS is a unitless measure of CAC highly correlated to the Agatston score but with numerical values assigned to individuals with Agatston score=0. We used linear mixed effect models to assess the association of baseline urinary metal levels with baseline SWCS, annual change in SWCS, and SWCS over ten years of follow-up. Urinary metals (adjusted to µg/g creatinine) and SWCS were log transformed. Models were progressively adjusted for baseline sociodemographic factors, estimated glomerular filtration rate, lifestyle factors, and clinical factors.

**Results:** At baseline, the median and interquartile range (25^th^, 75^th^) of SWCS was 6.3 (0.7, 58.2). For urinary cadmium, the fully adjusted geometric mean ratio (GMR) (95%Cl) of SWCS comparing the highest to the lowest quartile was 1.51 (1.32, 1.74) at baseline and 1.75 (1.47, 2.07) at ten years of follow-up. For urinary tungsten, uranium, and cobalt the corresponding GMRs at ten years of follow-up were 1.45 (1.23, 1.71), 1.39 (1.17, 1.64), and 1.47 (1.25, 1.74), respectively. For copper and zinc, the association was attenuated with adjustment for clinical risk factors; GMRs at ten years of follow-up before and after adjustment for clinical risk factors were 1.55 (1.30, 1.84) and 1.33 (1.12, 1.58), respectively, for copper and 1.85 (1.56, 2.19) and 1.57 (1.33, 1.85) for zinc.

**Conclusion:** Higher levels of cadmium, tungsten, uranium, cobalt, copper, and zinc, as measured in urine, were associated with subclinical CVD at baseline and at follow-up. These findings support the hypothesis that metals are pro-atherogenic factors.

**CLINICAL PERSPECTIVE:** What is new?

- Urinary levels of non-essential (cadmium, tungsten, uranium) and essential metals (cobalt, copper, zinc) are associated with coronary artery calcification at baseline and at ten years of follow up in a diverse US sample.

What are the clinical implications?

- Reductions in environmental metal exposure may improve cardiovascular health.
- Dietary and chelation interventions to reduce metals in the body may improve CVD outcomes.

## INTRODUCTION

Metals are ubiquitous contaminants that affect communities globally.^1^ In 2023, supported by epidemiologic and experimental evidence, the American Heart Association established lead, cadmium, and arsenic as cardiovascular disease (CVD) risk factors.^2^ Other metals may also promote atherosclerosis,^3, 4^ an inflammatory process underlying the most common forms of CVD. In the coronary arteries, atherosclerosis induces calcification, which can be measured non-invasively using the Agatston scoring method. Coronary artery calcification (CAC) is highly predictive of coronary heart disease events.^5^ Few studies have investigated the association of metals with CAC, therefore, the role of calcification in metal-related CVD is currently unknown.

Metals arise from anthropogenic and natural sources and vary geographically. Some are essential while others have no function in humans. Likewise, ambient particulate matter of diameter ≤2.5 µg/m^3^ (PM_2.5_) is an established risk factor for calcification,^6^ and may be composed of toxic metals.^7^ Metals differ in redox activity and, thus, on the potential toxicity mechanisms.^8^ Cobalt and copper, both essential elements, are examples of redox active metals capable of directly inducing reactive oxygen species, a precursor to the development of CVD.^9^ Conversely, the non-essential metal cadmium binds sulfhydryl groups and depletes glutathione, a protective antioxidant.^10^ Several metals additionally disrupt the endocrine system^11^ and target the vascular system,^12^ supporting that metals are atherogenic through multiple pathways.

The main objective of this study is to investigate the longitudinal association of urinary metal levels, biomarkers of metal exposure and internal dose, with changes in spatially weighted calcium scores (SWCS), a measure of CAC that has the advantage of providing numerical scores for individuals with Agatston scores equal to zero^13^ in a multi-ethnic and geographically diverse longitudinal study of adults in the US. We prioritized non-essential (cadmium, tungsten, uranium) and essential (cobalt, copper, zinc) metals that are relevant in US populations and have been previously associated with CVD outcomes.^3, 4^ Other metals that are difficult to interpret in urine (e.g., lead) or in populations with high levels of seafood intake (e.g., arsenic) or for which there is limited evidence of an association with CVD outcomes (e.g., cesium, strontium, manganese) were reported as secondary analyses.

## METHODS

### Study Population

The Multi-Ethnic Study of Atherosclerosis (MESA) is a multi-center, prospective cohort study of subclinical to clinical CVD.^14^ Between July 2000 and August 2002, MESA recruited 6,814 participants using community-based strategies at six study sites in Baltimore MD, Chicago IL, Los Angeles CA, New York NY, St. Paul MN, and Winston Salem NC. Participants were free of clinical CVD, men and women aged 45–84 years old from four race and ethnic groups (White, Black, Hispanic/Latino, and Chinese). Data were analyzed for follow-up through MESA Exam 5. Participants completed up to 5 clinic visits (Exam 1 in 2000-2002, Exam 2 in 2002-2004, Exam 3 in 2004-2006, Exam 4 in 2005-2007, and Exam 5 in 2010-2012) and 14 follow-up phone calls. All participants gave written informed consent. The Institutional Review Board at each study site approved the study.

Of the 6,814 MESA participants at baseline, 6,729 had metals and creatinine measured in urine at baseline (Figure S1). We excluded 4 participants with extreme metal values (3 observations for Co, 1 for Cu, 2 for U), as the levels for these participants were 100 times higher than the other highest values in the study. We excluded 32 participants who had a coronary revascularization procedure after exam 1 due to CAC measurement interference and 27 participants missing SWCS. Additionally, we excluded 21 participants missing data on education, 69 missing cigarette pack years, 2 missing physical activity, 98 missing low-density lipoprotein cholesterol (LDL), 4 missing diabetes status, 2 missing systolic blood pressure, 38 missing estimated glomerular filtration rate (eGFR), and 14 missing lipid lowering and blood pressure medications. The final sample size included a total of 6,418 participants with one or more repeated measures of SWCS for 15,643 observations, including 6,206 at baseline. Approximately 10% of participants (n=950) also had metals measured at Exam 5. Among the 6,418 participants in the study after removing missing data, 594 had urinary metal measures available at Exams 1 and 5.

### Urinary Metals

Spot urine samples were collected during mid to late morning at baseline Exams 1 and 5 using urine cups, aliquoted in small vials, shipped frozen on dry ice to the MESA biorepository, and stored at −80°C. In 2019, aliquots of 0.8mL urine were shipped on dry ice to the Trace Metals Core Laboratory at Columbia University. Detailed information on the analytical protocol to measure metals in MESA have been described elsewhere.^15^ Briefly, all metals were analyzed using PerkinElmer NexION 350S Inductively Coupled Plasma Mass Spectrometry with dynamic reaction cell (ICP-DRC-MS) instrument.^16^ At least five multi-element standard solutions were used for instrument calibration. The same diluent used for urine samples was used for calibration standards. Metal concentrations of the calibration solutions were chosen to cover the expected ranges of urine analyte concentrations. Samples were analyzed, blinded to participants’ characteristics, along with sample preparation blanks, and commercially available certified urine reference materials with a broad range of metal concentrations. Approximately 10% of the samples were prepared and measured in duplicate to determine intra-precision, and ∼10% were prepared and measured on different days to determine inter-precision. The intra- and inter-assay coefficient of variation ranged from 2.5% for zinc to 14% for uranium, and from 5.8% for cadmium to 16% for uranium, respectively (Table S1). Samples below the method detection limit (MDL) were divided by the √2. In most urine samples (>95%), the measured elemental concentrations exceeded the MDL except for uranium (11%) and tungsten (32%), see Table S1. To correct for urine dilution, we divided metal concentration by urine creatinine concentration (µg/creatinine), measured using the Jaffe reaction method.^17^ For participants with metals analyzed at Exams 1 and 5 (n=594), the intraclass correlation coefficient ranged from 0.50 to 0.72 for cobalt and uranium, respectively, supporting that a single baseline metal measure is a good reflection of long-term metal levels.

### Computed Tomography (cardiac CT) Scanning and Coronary Artery Calcification Measurement

All participants received cardiac CT scans at baseline to measure CAC as previously described.^18^ Scans were repeated for nearly all participants between 2002 and 2005, for a subset of participants between 2005 and 2007, and for half of all participants between 2010 and 2012. After arterial trajectories across the surface of the heart were determined within 8 mm, and a phantom-based adjustment was applied, candidate calcified plaques were identified by the software with the criteria that each plaque be composed of at least 4 contiguous voxels with an attenuation level of 130 Hounsfield units or greater. A radiologist or cardiologist scored all CT scans using an interactive scoring system at the Harbor-UCLA Los Angeles Biomedical Research Institute by the Agatston method.^18^ The Agatston score (AS) reproducibly quantitates CAC from CT images and is highly predictive of coronary heart disease (CHD) and CVD events.^19^ CAC-AS is a continuous measure that is dichotomized as 0 and 1 or higher, respectively, for any calcification below or above the threshold.

### Spatially Weighted Calcium Score

The traditional Agatston score ignores available information in the CT scan due to the conservative but specific algorithm for lesion detection.^13, 20^ Therefore, participants early in the calcification process who do not meet the traditional threshold for the presence of CAC on the CT scan are classified as having a CAC-AS=0. The spatially weighted calcium score (SWCS) is a semi-automated threshold-free CAC scoring method. As described previously,^13^ weights were assigned to each image voxel to calibrate and weight according to the phantom to maximize the CT scan information. Each voxel was assigned a score dependent on the voxel weight and neighboring voxel weight. The detailed algorithm for calculating the SWCS is published.^13^ SWCS is a continuous measure of calcification that provides a quantifiable CAC level even when CAC-AS=0, and that is very similar to CAC-AS when it is >0. SWCS predicts incident CHD events even among participants with CAC-AS=0, supporting it is an excellent marker of atherosclerotic CVD risk even at low levels of coronary calcification.^20^

### Covariates

Age, sex, race and ethnicity, education, smoking status, physical activity, and use of lipid-lowering and hypertension medications were collected by questionnaire during Exam 1. Race and ethnicity were self-reported and categorized as White, Black, Hispanic/Latino, and Chinese. Study sites included Baltimore MD, Chicago IL, Los Angeles CA, New York NY, St. Paul MN, and Winston Salem NC. Cigarette smoking status was classified as never, former, and current. Participants who had not smoked 100 cigarettes in their lifetime were classified as never smokers. Participants who answered yes were classified as current smokers if they had smoked in the last 30 days or classified as former smokers if they had not smoked in the last 30 days. Cigarette pack-years was calculated by multiplying the intensity in packs per day by duration in years where 20 cigarettes define a pack. Physical activity was defined as the total moderate and high physical activity in hours per week, Monday to Sunday.

At Exam 1, height and weight were measured to calculate body mass index (BMI, kg/m^2^). Resting systolic and diastolic blood pressure were measured three times in the seated position using a Dinamap model Pro 100 automated oscillometric sphygmomanometer with the last two measurements averaged for analysis. Low- and high-density lipoprotein cholesterol (LDL, HDL, mg/dL blood), and calibrated fasting plasma glucose (FPG, mg/dL blood), were assessed using standard laboratory techniques. Diabetes mellitus (DM) was defined by the 2003 American Diabetes Association fasting criteria and categorized by normal (<100 mg/dL blood FPG), impaired fasting glucose (100-125 mg/dL blood FPG), untreated and treated diabetes (≥126 mg/dL blood FPG or taking diabetes medications). eGFR was calculated using the new creatinine and cyastin-C based Chronic Kidney Disease Epidemiology Collaboration equation without accounting for race and ethnicity.^21^ eGFR can influence metal excretion in urine and was therefore used for adjustment in our models. Urinary cotinine, a metabolite of nicotine, was measured by immune assay (Immulite 2000 Nicotine Assay; Diagnostic products Corp., Los Angeles, CA) in a subset of participants (n=3,791). Finally, because air pollution is a source of metal exposure,^7^ average PM_2.5_ (µg/m^3^) was estimated using predictions from city-specific spatiotemporal models for calendar years 2000-2001 at baseline.^22^

### Statistical Analysis

We conducted descriptive analyses overall and by participant characteristics of continuous SWCS, dichotomous CAC-AS, and urinary metal levels. Urinary metal levels and SWCS were right skewed and log-transformed for analysis. We performed Spearman correlation tests for log-transformed urinary metals (µg/g creatinine).

We used mixed effect models on log-transformed repeated SWCS measures by baseline urinary metal levels with a random intercept on the participant and random slope on the time since baseline cardiac CT scan. By exponentiating the coefficients, the model allows to estimate baseline geometric mean ratios (GMRs), annual GMR change, and GMRs at a relevant time during the follow-up (we selected 10 years) in the average person. Urinary metal levels were modeled as: (1) per interquartile range (IQR) on log-transformed levels (to compare the 75^th^ to the 25^th^ percentile), (2) quartiles (to compare each of the highest three to the lowest quartiles), and (3) log-transformed concentrations with restricted quadratic splines (to evaluate the flexible dose-response relationship). We evaluated the association of baseline metal levels with dichotomous CAC-AS score using a modified Poisson with the generalized linear mixed model to estimate relative risk of incident CAC-AS>0 among participant with CAC-AS=0 at baseline.

Model 1 was adjusted for sociodemographic (age, sex, race and ethnicity, study site, education) and behavioral factors (smoking status, pack-years, physical activity) and eGFR and BMI. Model 2 was additionally adjusted for cardiovascular risk factors (systolic blood pressure, antihypertensive medications, LDL-cholesterol, HDL-cholesterol, lipid lowering medications, and diabetes status). Because urinary metals levels were measured at baseline, all adjustments were time-invariant covariates acquired at baseline. For the dose-response figures we only show the results for model 2. Finally, we used Wald tests and conducted subgroup analysis to assess effect modification by subgroups of age, sex, race and ethnicity, smoking status, and diabetes status for geometric mean ratios at baseline and at ten years of follow-up.

### Sensitivity Analyses

We conducted several sensitivity analyses. Because SWCS showed a potential nonlinear relationship with time over the follow-up, we modeled the time since last CT scan as a polynomial. The effect estimates remained unchanged and thus was not used in our final models (not shown). We further adjusted for city-specific average PM_2.5_ at baseline to account for metal exposures originating from ambient air pollution and for potential confounding of the relationship to CAC.^6^ We also further adjusted our models for urinary cotinine to determine whether tobacco use, use of an unaccounted-for nicotine product, or secondhand tobacco exposure is accurately captured by self-reported data. Urinary creatinine levels are commonly used to account for urine dilution but vary by age, sex, and other characteristics. We conducted a sensitivity analysis with adjustment for urine specific gravity instead of using urinary creatinine. Because diabetes status can impact urinary zinc levels, we further adjusted for fasting plasma glucose levels. Finally, in a small subset of participants (n=594), we investigated the relationship between time varying urinary metal levels at two time points, Exams 1 and 5, with repeated measures of SWCS.

## RESULTS

The median and interquartile ranges (IQR) [25^th^, 75^th^] of SWCS was 6.3 (0.7, 58.2) and CAC-AS>0 occurred in approximately 50% participants at baseline (Table 1). Median and IQR [25^th^, 75^th^] of SWCS and frequency of positive CAC-AS increased with age and were higher among males, White participants, and those with a high school education or less. Participants who formerly smoked and those who had diabetes mellitus and hypertension had higher median SWCS and frequency of positive CAC-AS.

**Table 1.** Median and interquartile ranges [25^th^, 75^th^] of spatially weighted calcium scores (SWCS) and corresponding number (%) of positive coronary artery calcification Agatston scores (CAC-AS) overall and by participant characteristic at baseline exam 1 (2000-2002).

|  | **n** | **SWCS** | **CAC-AS ≥ 1** |
| --- | --- | --- | --- |
| Overall | 6,206 | 6.3 [0.7, 58.2] | 3,080 (49.6) |
| Age (Years) |  |  |  |
| <55 | 1,758 | 1.5 [0.3, 7.1] | 429 (24.4) |
| 55-64 | 1,716 | 4.4 [0.6, 31.7] | 759 (44.2) |
| ≥65 | 2,732 | 30.2 [2.9, 162.4] | 1,892 (69.3) |
| Sex (%) |  |  |  |
| Female | 3,284 | 2.9 [0.4, 23.8] | 1,302 (39.6) |
| Male | 2,922 | 17.1 [2, 117.9] | 1,778 (60.8) |
| Race and Ethnicity (%) |  |  |  |
| White | 2,375 | 8.8 [0.7, 102.3] | 1,349 (56.8) |
| Black | 1,706 | 5.6 [0.8, 36.2] | 741 (43.4) |
| Hispanic/Latino | 1,360 | 6 [1.4, 45] | 609 (44.8) |
| Chinese | 765 | 3.1 [0.3, 45.9] | 381 (49.8) |
| Study Site (%) |  |  |  |
| Salem, NC | 960 | 2.1 [0.1, 50] | 489 (50.9) |
| New York, NY | 994 | 6.7 [1.3, 35.7] | 412 (41.4) |
| Baltimore, MD | 955 | 10.2 [1.3, 89.3] | 532 (55.7) |
| St. Paul, MN | 972 | 5.7 [1.6, 76.4] | 496 (51) |
| Chicago, IL | 1,112 | 6.2 [0.5, 52.4] | 534 (48) |
| Los Angeles, CA | 1,213 | 8.7 [0.8, 56.8] | 617 (50.9) |
| Education (%) |  |  |  |
| High School or Less | 2,226 | 8.1 [1.2, 65.9] | 1,169 (52.5) |
| Some College | 1,450 | 6.5 [0.8, 57.8] | 702 (48.4) |
| College Degree or More | 2,530 | 4.8 [0.5, 49.7] | 1,209 (47.8) |
| Smoking Status (%) |  |  |  |
| Never | 3,165 | 4.3 [0.5, 37.4] | 1,400 (44.2) |
| Former | 2,248 | 12.1 [1.3, 92.4] | 1,296 (57.7) |
| Current | 793 | 4.7 [0.7, 51.9] | 384 (48.4) |
| Pack-Years |  |  |  |
| 0 | 3,250 | 4.3 [0.5, 37.1] | 1,431 (44) |
| 1 - 10 | 1,084 | 5.8 [0.9, 52.5] | 515 (47.5) |
| 11 - 20 | 604 | 8.9 [1, 63.7] | 322 (53.3) |
| >20 | 1,268 | 18.3 [1.5, 141.8] | 812 (64) |
| BMI (kg/ |  |  |  |
| BMI < 25 | 1,810 | 2.2 [0.2, 51.2] | 862 (47.6) |
| 25 ≤ BMI < 30 | 2,432 | 5.6 [0.8, 62.2] | 1,252 (51.5) |
| BMI ≥ 30 | 1,964 | 11.3 [2.3, 58.8] | 966 (49.2) |
| Physical Activity (MET-hours/week) |  |  |  |
| ≤34 | 1,625 | 9 [0.9, 62.9] | 846 (52.1) |
| 35-69 | 1,563 | 6.9 [0.7, 65.5] | 816 (52.2) |
| 70-139 | 1,691 | 6.1 [0.7, 60.4] | 830 (49.1) |
| ≥140 | 1,327 | 4.5 [0.6, 37] | 588 (44.3) |
| LDL-cholesterol (mg/dL) |  |  |  |
| <100 | 1,794 | 5.6 [0.6, 60] | 865 (48.2) |
| 100-129 | 2,414 | 6.5 [0.7, 56.3] | 1,183 (49) |
| 130-159 | 1,435 | 6.3 [0.8, 58.6] | 725 (50.5) |
| ≥160 | 563 | 7.3 [1.1, 62.2] | 307 (54.5) |
| HDL-cholesterol (mg/dL) |  |  |  |
| <50 | 3,238 | 10 [1.3, 71.2] | 1,750 (54) |
| 50-69 | 2,287 | 4.5 [0.5, 43.3] | 1,047 (45.8) |
| 70-99 | 625 | 2 [0.2, 31.5] | 256 (41) |
| ≥100 | 56 | 3.8 [0.3, 51.6] | 27 (48.2) |
| eGFR (mL/min/1.73 m<sup>2</sup>) |  |  |  |
| <45 | 55 | 81.5 [20, 230.6] | 44 (80) |
| 45-59 | 140 | 55.8 [4.9, 293.2] | 107 (76.4) |
| 60-89 | 1,739 | 23.1 [2.6, 134.8] | 1,141 (65.6) |
| >90 | 4,272 | 3.6 [0.5, 30.1] | 1,788 (41.9) |
| Diabetes Mellitus (%) |  |  |  |
| Normal | 4,602 | 4.3 [0.5, 43.3] | 2,124 (46.2) |
| IFG | 849 | 12.6 [1.9, 84.1] | 483 (56.9) |
| DM | 755 | 22.1 [3.4, 151.6] | 473 (62.6) |
| Hypertension (%) |  |  |  |
| No | 3,415 | 3 [0.4, 27.2] | 1,398 (40.9) |
| Yes | 2,791 | 17.2 [1.9, 110.5] | 1,682 (60.3) |
| BP Medications (%) |  |  |  |
| No | 3,897 | 3.6 [0.5, 32.6] | 1,681 (43.1) |
| Yes | 2,309 | 18.1 [2, 113.7] | 1,399 (60.6) |
| Lipid Medications (%) |  |  |  |
| No | 5,186 | 5 [0.6, 45.8] | 2,400 (46.3) |
| Yes | 1,020 | 23 [2, 144.7] | 680 (66.7) |
| PM<sub>2.5</sub> (μg/m<sup>3</sup>) |  |  |  |
| ≤12 | 913 | 6.1 [1.6, 83.3] | 473 (51.8) |
| 13-15 | 264 | 5.5 [0.6, 45.6] | 125 (47.3) |
| 16-20 | 3,707 | 6.3 [0.6, 57.4] | 1,820 (49.1) |
| >20 | 1,063 | 8.7 [0.8, 57.2] | 547 (51.5) |

Non-essential and essential urinary metal levels varied by participant characteristic (Figure 1). Urinary metal levels (µg/g creatinine) tended to be higher among females, older participants, Chinese participants, and those with less education. Participants from Los Angeles had markedly higher urinary tungsten and uranium levels, and somewhat higher cadmium, cobalt, and copper levels. Cadmium levels were higher among current smokers; the essential metals cobalt and copper were lower among current smokers. Spearman correlation values of urinary metal levels ranged between 0.01 and 0.61 (Figure S2).

**Figure 1.**
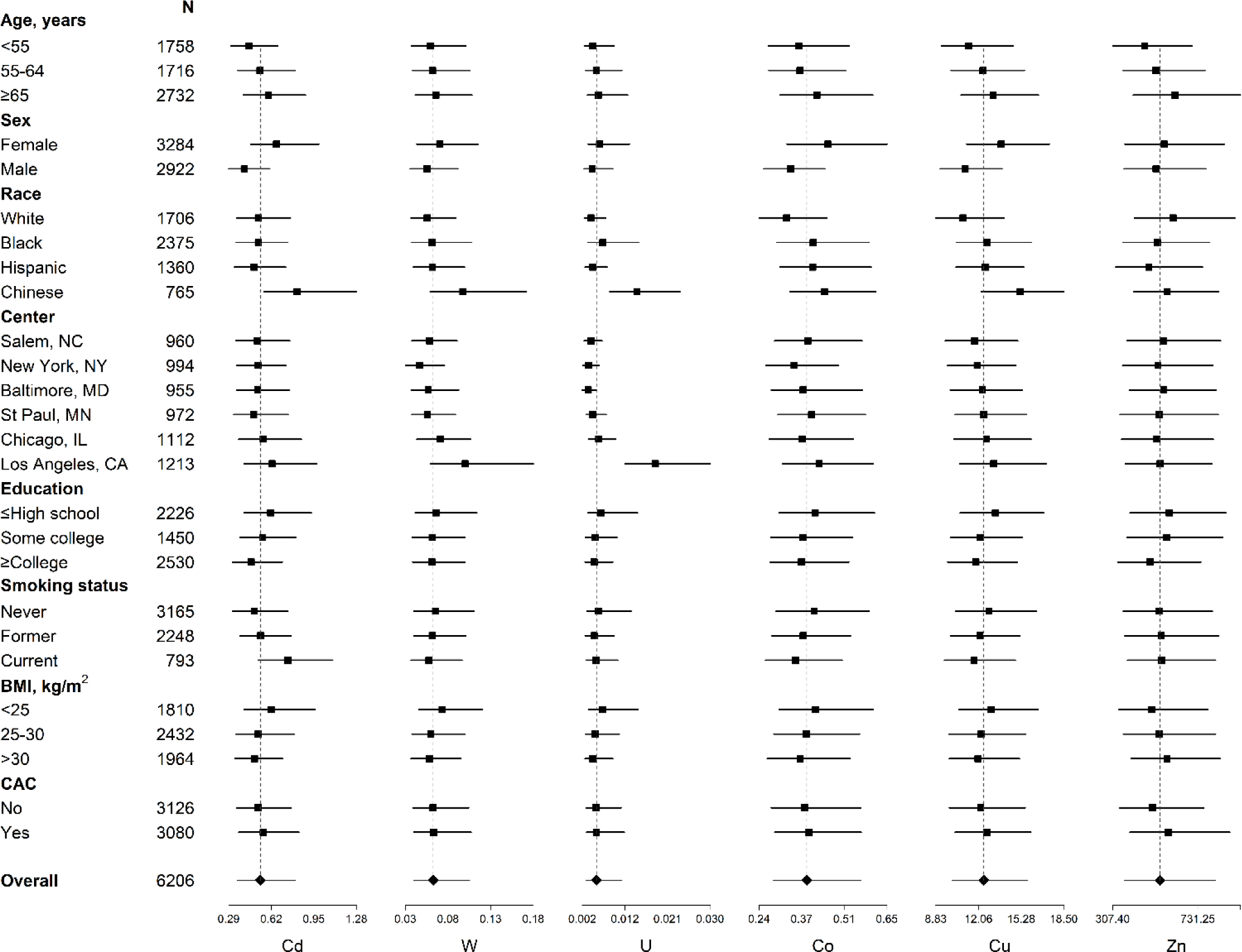
Median and interquartile ranges [25^th^, 75^th^] of urine metal levels (µg/g creatinine) by participant characteristic for non-essential metals cadmium (Cd), tungsten (W), and uranium (U), and essential metals cobalt (Co), copper (Cu), and zinc (Zn). Points represent the median urine metal level and lines correspond to the interquartile range overall and for each subgroup at baseline. The n for each group is on the y-axis. The dotted line represents the overall median urine metal level.

The fully adjusted GMR (95%CI) of SWCS comparing the highest to lowest urinary cadmium quartile was 1.51 (1.32, 1.74) at baseline and 1.75 (1.47, 2.07) at ten years of follow-up; the annual change was positive but not statistically significant (Table 2). The non-linear association apparent in the quartile models was also observed with the restricted quadratic spline models, with clear positive dose-response relationships with SWCS observed for urinary cadmium above 0.5 µg/g creatinine both at baseline and at ten years of follow-up (Figure 2).

**Figure 2.**
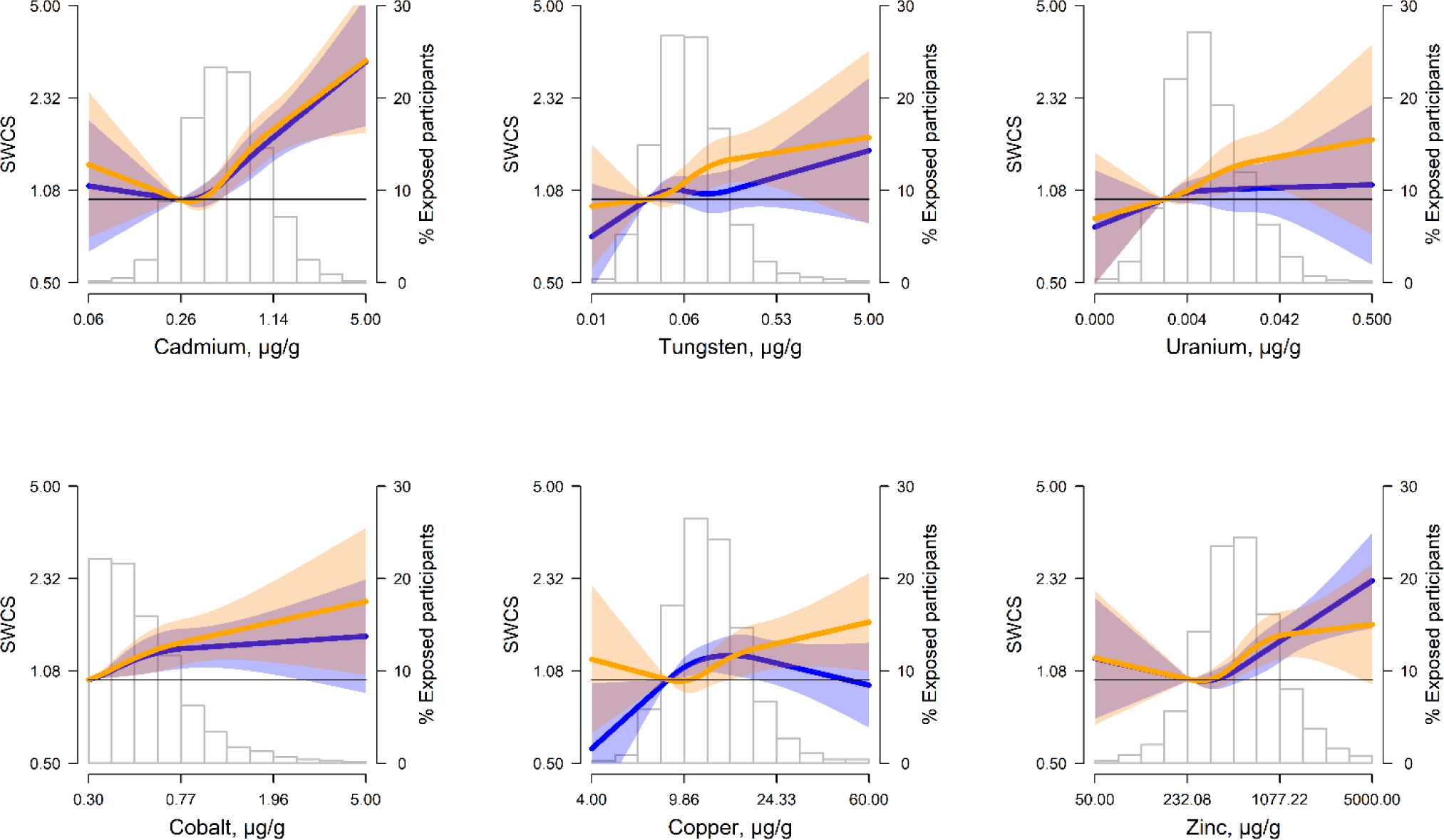
Geometric mean ratio (GMR) (95% confidence interval) of spatially weighted calcium scores (SWCS) at baseline (blue lines and shaded areas) and at 10-years of follow-up (orange lines and shaded areas) by urinary metal levels (µg/g creatinine) modeled as restricted cubic splines. Lines (shaded areas) represent the GMR (95%CI) of SWCS by metals modeled as restricted cubic splines for log transformed metal distributions with knots at 10^th^, 50^th^, and 90^th^ percentiles. The reference value was set at the 10^th^ percentile. Models were adjusted for age, sex, race and ethnicity, study site, education, eGFR, smoking status, pack-years, physical activity, BMI, systolic blood pressure, antihypertensive medication, LDL-cholesterol, HDL-cholesterol, lipid lowering medications, and diabetes status. The histograms in the background represent the distribution of each metal (µg/g creatinine) at baseline.

**Table 2.**
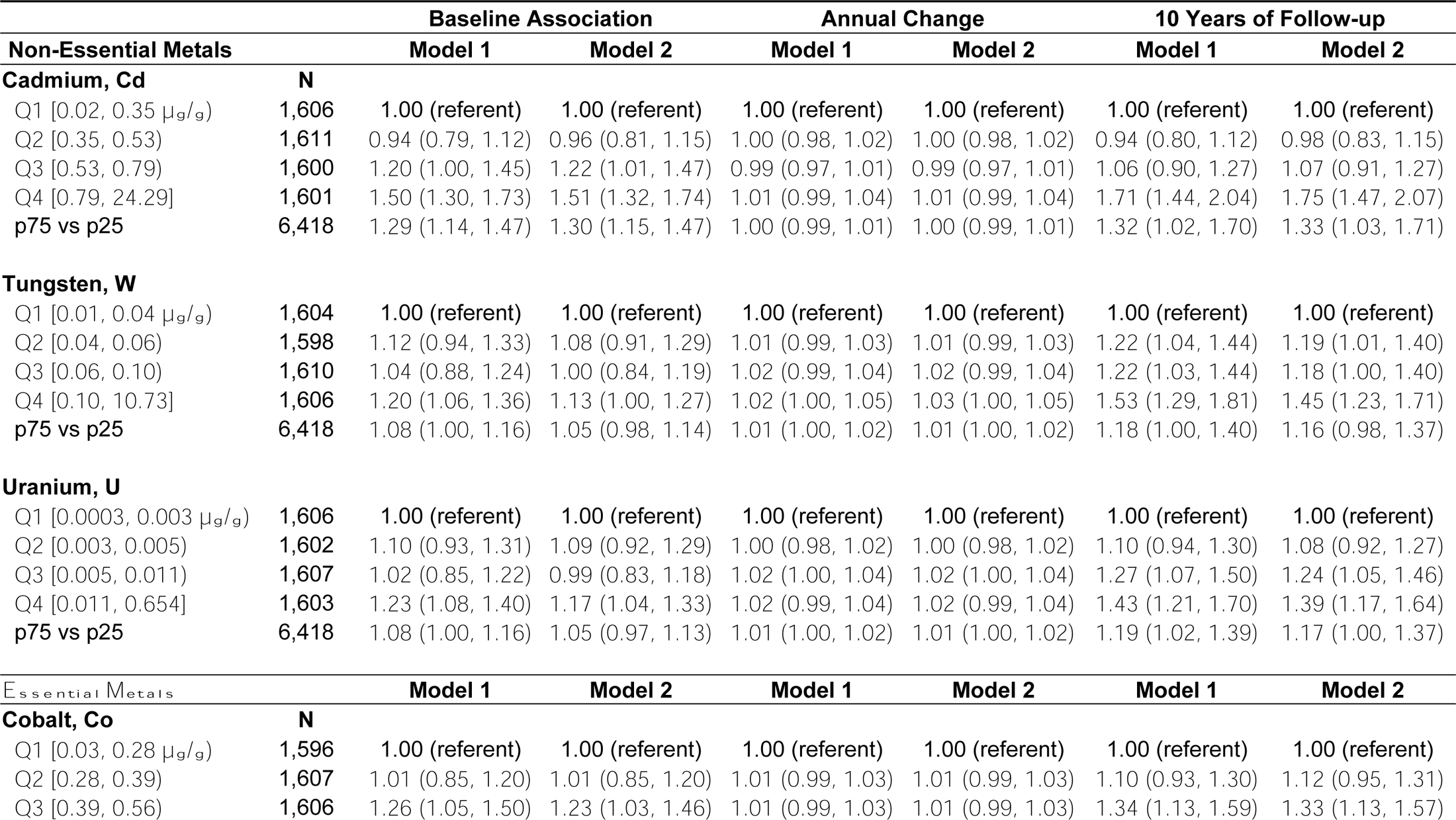
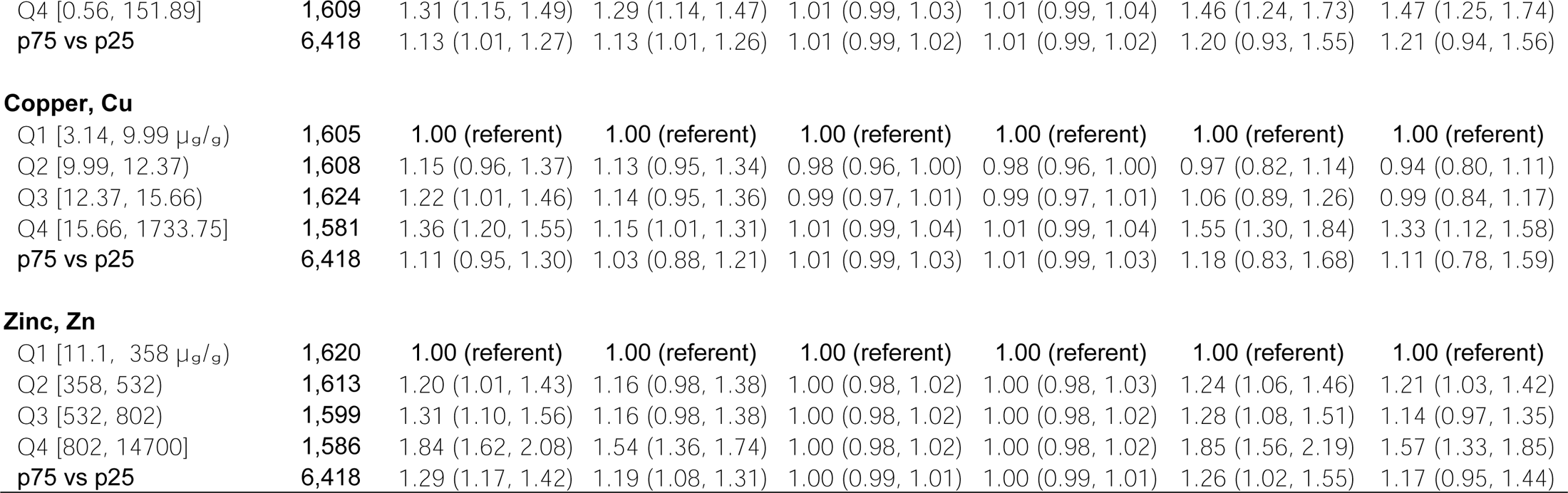
Geometric mean ratio (GMR) (95% confidence interval) of spatially weighted calcium scores (SWCS) by levels (µg/g creatinine) of non-essential and essential metals in urine. Model 1 was adjusted for age, sex, race and ethnicity, study site, education, eGFR, smoking status, pack-years, physical activity and BMI. Model 2 was additionally adjusted for systolic blood pressure, antihypertensive medication, LDL-cholesterol, HDL-cholesterol, lipid lowering medications, and diabetes status.

|  |  | Baseline Association |  | Annual Change |  | 10 Years of Follow-up |  |
| --- | --- | --- | --- | --- | --- | --- | --- |
| Non-Essential Metals |  | Model 1 | Model 2 | Model 1 | Model 2 | Model 1 | Model 2 |
| **Cadmium, Cd** | **N** |  |  |  |  |  |  |
| Q1 [0.02, 0.35 µg/g) | 1,606 | 1.00 (referent) | 1.00 (referent) | 1.00 (referent) | 1.00 (referent) | 1.00 (referent) | 1.00 (referent) |
| Q2 [0.35, 0.53) | 1,611 | 0.94 (0.79, 1.12) | 0.96 (0.81, 1.15) | 1.00 (0.98, 1.02) | 1.00 (0.98, 1.02) | 0.94 (0.80, 1.12) | 0.98 (0.83, 1.15) |
| Q3 [0.53, 0.79) | 1,600 | 1.20 (1.00, 1.45) | 1.22 (1.01, 1.47) | 0.99 (0.97, 1.01) | 0.99 (0.97, 1.01) | 1.06 (0.90, 1.27) | 1.07 (0.91, 1.27) |
| Q4 [0.79, 24.29] | 1,601 | 1.50 (1.30, 1.73) | 1.51 (1.32, 1.74) | 1.01 (0.99, 1.04) | 1.01 (0.99, 1.04) | 1.71 (1.44, 2.04) | 1.75 (1.47, 2.07) |
| p75 vs p25 | 6,418 | 1.29 (1.14, 1.47) | 1.30 (1.15, 1.47) | 1.00 (0.99, 1.01) | 1.00 (0.99, 1.01) | 1.32 (1.02, 1.70) | 1.33 (1.03, 1.71) |
| **Tungsten, W** |  |  |  |  |  |  |  |
| Q1 [0.01, 0.04 µg/g) | 1,604 | 1.00 (referent) | 1.00 (referent) | 1.00 (referent) | 1.00 (referent) | 1.00 (referent) | 1.00 (referent) |
| Q2 [0.04, 0.06) | 1,598 | 1.12 (0.94, 1.33) | 1.08 (0.91, 1.29) | 1.01 (0.99, 1.03) | 1.01 (0.99, 1.03) | 1.22 (1.04, 1.44) | 1.19 (1.01, 1.40) |
| Q3 [0.06, 0.10) | 1,610 | 1.04 (0.88, 1.24) | 1.00 (0.84, 1.19) | 1.02 (0.99, 1.04) | 1.02 (0.99, 1.04) | 1.22 (1.03, 1.44) | 1.18 (1.00, 1.40) |
| Q4 [0.10, 10.73] | 1,606 | 1.20 (1.06, 1.36) | 1.13 (1.00, 1.27) | 1.02 (1.00, 1.05) | 1.03 (1.00, 1.05) | 1.53 (1.29, 1.81) | 1.45 (1.23, 1.71) |
| p75 vs p25 | 6,418 | 1.08 (1.00, 1.16) | 1.05 (0.98, 1.14) | 1.01 (1.00, 1.02) | 1.01 (1.00, 1.02) | 1.18 (1.00, 1.40) | 1.16 (0.98, 1.37) |
| **Uranium, U** |  |  |  |  |  |  |  |
| Q1 [0.0003, 0.003 µg/g) | 1,606 | 1.00 (referent) | 1.00 (referent) | 1.00 (referent) | 1.00 (referent) | 1.00 (referent) | 1.00 (referent) |
| Q2 [0.003, 0.005) | 1,602 | 1.10 (0.93, 1.31) | 1.09 (0.92, 1.29) | 1.00 (0.98, 1.02) | 1.00 (0.98, 1.02) | 1.10 (0.94, 1.30) | 1.08 (0.92, 1.27) |
| Q3 [0.005, 0.011) | 1,607 | 1.02 (0.85, 1.22) | 0.99 (0.83, 1.18) | 1.02 (1.00, 1.04) | 1.02 (1.00, 1.04) | 1.27 (1.07, 1.50) | 1.24 (1.05, 1.46) |
| Q4 [0.011, 0.654] | 1,603 | 1.23 (1.08, 1.40) | 1.17 (1.04, 1.33) | 1.02 (0.99, 1.04) | 1.02 (0.99, 1.04) | 1.43 (1.21, 1.70) | 1.39 (1.17, 1.64) |
| p75 vs p25 | 6,418 | 1.08 (1.00, 1.16) | 1.05 (0.97, 1.13) | 1.01 (1.00, 1.02) | 1.01 (1.00, 1.02) | 1.19 (1.02, 1.39) | 1.17 (1.00, 1.37) |
| Essential Metals |  | Model 1 | Model 2 | Model 1 | Model 2 | Model 1 | Model 2 |
| **Cobalt, Co** | **N** |  |  |  |  |  |  |
| Q1 [0.03, 0.28 µg/g) | 1,596 | 1.00 (referent) | 1.00 (referent) | 1.00 (referent) | 1.00 (referent) | 1.00 (referent) | 1.00 (referent) |
| Q2 [0.28, 0.39) | 1,607 | 1.01 (0.85, 1.20) | 1.01 (0.85, 1.20) | 1.01 (0.99, 1.03) | 1.01 (0.99, 1.03) | 1.10 (0.93, 1.30) | 1.12 (0.95, 1.31) |
| Q3 [0.39, 0.56) | 1,606 | 1.26 (1.05, 1.50) | 1.23 (1.03, 1.46) | 1.01 (0.99, 1.03) | 1.01 (0.99, 1.03) | 1.34 (1.13, 1.59) | 1.33 (1.13, 1.57) |
| Q4 [0.56, 151.89] | 1,609 | 1.31 (1.15, 1.49) | 1.29 (1.14, 1.47) | 1.01 (0.99, 1.03) | 1.01 (0.99, 1.04) | 1.46 (1.24, 1.73) | 1.47 (1.25, 1.74) |
| p75 vs p25 | 6,418 | 1.13 (1.01, 1.27) | 1.13 (1.01, 1.26) | 1.01 (0.99, 1.02) | 1.01 (0.99, 1.02) | 1.20 (0.93, 1.55) | 1.21 (0.94, 1.56) |
| Copper, Cu |  |  |  |  |  |  |  |
| Q1 [3.14, 9.99 µg/g) | 1,605 | 1.00 (referent) | 1.00 (referent) | 1.00 (referent) | 1.00 (referent) | 1.00 (referent) | 1.00 (referent) |
| Q2 [9.99, 12.37) | 1,608 | 1.15 (0.96, 1.37) | 1.13 (0.95, 1.34) | 0.98 (0.96, 1.00) | 0.98 (0.96, 1.00) | 0.97 (0.82, 1.14) | 0.94 (0.80, 1.11) |
| Q3 [12.37, 15.66) | 1,624 | 1.22 (1.01, 1.46) | 1.14 (0.95, 1.36) | 0.99 (0.97, 1.01) | 0.99 (0.97, 1.01) | 1.06 (0.89, 1.26) | 0.99 (0.84, 1.17) |
| Q4 [15.66, 1733.75] | 1,581 | 1.36 (1.20, 1.55) | 1.15 (1.01, 1.31) | 1.01 (0.99, 1.04) | 1.01 (0.99, 1.04) | 1.55 (1.30, 1.84) | 1.33 (1.12, 1.58) |
| p75 vs p25 | 6,418 | 1.11 (0.95, 1.30) | 1.03 (0.88, 1.21) | 1.01 (0.99, 1.03) | 1.01 (0.99, 1.03) | 1.18 (0.83, 1.68) | 1.11 (0.78, 1.59) |
| Zinc, Zn |  |  |  |  |  |  |  |
| Q1 [11.1, 358 µg/g) | 1,620 | 1.00 (referent) | 1.00 (referent) | 1.00 (referent) | 1.00 (referent) | 1.00 (referent) | 1.00 (referent) |
| Q2 [358, 532) | 1,613 | 1.20 (1.01, 1.43) | 1.16 (0.98, 1.38) | 1.00 (0.98, 1.02) | 1.00 (0.98, 1.03) | 1.24 (1.06, 1.46) | 1.21 (1.03, 1.42) |
| Q3 [532, 802) | 1,599 | 1.31 (1.10, 1.56) | 1.16 (0.98, 1.38) | 1.00 (0.98, 1.02) | 1.00 (0.98, 1.02) | 1.28 (1.08, 1.51) | 1.14 (0.97, 1.35) |
| Q4 [802, 14700] | 1,586 | 1.84 (1.62, 2.08) | 1.54 (1.36, 1.74) | 1.00 (0.98, 1.02) | 1.00 (0.98, 1.02) | 1.85 (1.56, 2.19) | 1.57 (1.33, 1.85) |
| p75 vs p25 | 6,418 | 1.29 (1.17, 1.42) | 1.19 (1.08, 1.31) | 1.00 (0.99, 1.01) | 1.00 (0.99, 1.01) | 1.26 (1.02, 1.55) | 1.17 (0.95, 1.44) |

For tungsten and uranium, the fully adjusted GMRs (95%CI) of SWCS at baseline comparing the highest to lowest quartiles were 1.13 (1.00, 1.27) and 1.17 (1.04, 1.33); the corresponding GMRs for the annual change were 1.03 (1.00, 1.05) and 1.02 (0.99, 1.04), respectively, and at 10 years of follow-up, they were 1.45 (1.23, 1.71) for tungsten and 1.39 (1.17, 1.64) for uranium. The flexible spline models were consistent with a linear dose-response, in particular at ten years of follow-up.

The fully adjusted GMRs (95%CI) of SWCS at baseline and ten years of follow-up comparing the highest to lowest essential metal quartiles were 1.29 (1.14, 1.47) and 1.47 (1.25, 1.74) for cobalt, 1.15 (1.01, 1.31) and 1.33 (1.12, 1.58) for copper, and 1.54 (1.36, 1.74) and 1.57 (1.33, 1.85) for zinc. For the three essential metals, the association with the annual change was not significant (Table 2), and the dose-responses tended to be flat at lower levels and positive at higher levels, especially at ten years (Figure 2). For copper and zinc, there was a marked decline in the association with SWCS both at baseline and at ten years of follow-up after adjusting for clinical risk factors (model 2) compared to model 1. In a post-hoc analysis, this attenuation was largely due to adjustment for diabetes status and fasting plasma glucose (Figure S3), and not to the other variables.

We conducted several sensitivity analyses. Further adjustment of the association between urinary metals and SWCS for ambient PM_2.5_ resulted in similar effect estimates (Figure S4). Further adjustment for urinary cotinine attenuated the association between urinary cadmium and SWCS, although the association remained significant (Figure S5). Estimates of the association with SWCS were attenuated when urinary metals were adjusted by urinary specific gravity instead of dividing by urinary creatinine, but the general patterns remained (Figure S6).

Using incident CAC-AS>0 over the follow-up as the study outcome, resulted in consistent associations of non-essential (cadmium, tungsten, uranium) and essential (cobalt, copper, zinc) metals with CAC compared to the SWCS models (Table S2). Models 1 and 2 for the association with SWCS of other non-essential (arsenic, barium, cesium, lead, strontium, thallium) and essential (manganese, molybdenum, selenium) elements available in MESA are reported in Table S3.

In stratified models by participant subgroups for the priority metals (Table S4), the associations remained similar by age group both at baseline and 10 years for all the metals and no consistent patterns were observed by race and ethnicity with differences for the same metals between baseline and follow-up. By sex the association for cadmium was stronger in women both at baseline and follow-up (p-value for interaction only significant at baseline), while for the other metals the patterns were inconsistent. By smoking status, the association for cadmium and uranium were stronger for former smokers at baseline and follow-up; patterns for other metals were inconsistent.

Finally, in the small subset of participants with exposure measures at two time points (n=594), the effect estimates were significant and even stronger compared to the main models based on Exam 1 data for urinary cadmium (Table S5), consistent but not significant for tungsten and uranium at ten years of follow-up and for copper at baseline and ten years of follow-up, and inconsistent but not significant for cobalt and zinc.

## DISCUSSION

In this longitudinal study of coronary atherosclerosis progression among multi-ethnic adults from six urban areas in the United States, we found that baseline urinary levels of the non-essential metals: cadmium, uranium, and tungsten, and essential metals: cobalt, copper, and zinc were associated with CAC, an established subclinical marker of CVD risk, at ten-years of follow-up. Among the non-essential metals, tungsten and uranium were significantly associated with annual changes in SWCS and with stronger associations at the ten years follow-up compared to baseline, while for cadmium the association at baseline and the 10 years follow-up remained similar. For copper and zinc, a marked attenuation of the association with SWCS was observed after adjustment for clinical risk factors, in particular diabetes and fasting plasma glucose. Taken together, these results support that metal exposure and/or metabolism, as measured in urine, contributes to the progression of atherosclerosis as measured by coronary calcification in diverse adults across the US.

### Non-essential Metals

#### Cadmium, Cd

Cadmium is a highly toxic and carcinogenic metal that has been associated with clinical CVD outcomes in numerous studies.^2, 23, 24^ In MESA, higher levels of urinary cadmium were associated with higher SWCS both at baseline and after ten-years of follow-up. The association with annual changes in SWCS, although positive, was not statistically significant. These findings are potentially related to the long-half life and thus urinary cadmium, which reflects the cumulative body burden. A meta-analysis of 12 prospective studies comparing the highest to lowest cadmium exposure categories and clinical CVD reported pooled relative risk of 1.36 (95% CI: 1.11-1.66) for urine.^25^ Proposed mechanisms by which cadmium affects the vasculature include impaired nitric oxide functioning and signaling,^26^ modulated calcium concentrations,^27^ endothelial cell apoptosis,^28^ and oxidative stress through glutathione depletion and other mechanisms.^29^ In Swedish adults (n=5,627), blood cadmium levels were cross-sectionally associated with the prevalence of CAC-AS>0 (prevalence ratio 1.25, 95% CI: 1.13, 1.38).^30^ Although results were similar, our study uses urinary cadmium, which has a longer half-life than blood cadmium. Tobacco smoke is the main source of cadmium followed by contaminated foods due to widespread soil pollution from the use of phosphate fertilizers rich in cadmium, production and disposal of nickel-cadmium batteries, and other industrial uses.^31^ We found consistent, although attenuated associatons between urinary cadmium and SWCS in never smokers in MESA, which is consistent with findings for the association with incident CVD among never smokers in the above cited meta-analysis (pooled relative risk 1.27, 95% CI: 0.97, 1.67).

#### Tungsten, W

Tungsten is widedspread in drinking water in the Western United States, used in welding, oil production, and electrical and aerospace industries, and exists in the particulate phase in ambient air due to low vapor pressure.^32^ Urinary tungsten levels have been associated with PM_2.5_ previously in MESA.^33^ Some prior but limited evidence has related tungsten with cardiovascular outcomes. In NHANES, urinary tungsten levels were associated with stroke prevalence,^34^ composite cardiovascular and cerebrovascular disease,^35^ and higher self-reported CVD.^36^ In the Strong Heart Study, increasing baseline urinary tungsten was not associated with incident CVD (n=2,726). ^37^ Our sensitivity analyses found a slight attenuation on the association of tungsten with SWCS at follow-up when further adjusting for PM_2.5._ Together these findings support that urinary tungsten levels contribute to the progression of atherosclerosis and that ambient air pollution may be a relevant source of tungsten. Additional studies are needed to evaluate the role of tungsten in atherosclerosis and clinical CVD, including relevant modifiers and confounders like PM_2.5._

#### Uranium, U

Uranium is present in groundwater, is used for nuclear energy production, and is often found in phosphate fertilizers due to uranium in the phosphate rock used for fertilizer manufacturing.^38^ Uranium exposure likely comes from groundwater contamination, which is federally regulated at a maximum contaminant level of 30 µg/L in drinking water. Previous experimental and epidemiological evidence of uranium exposure and CVD is limited.^8^ Three studies of enriched uranium miners found increased risk for CHD risk at three recruitment sites in the United States,^39^ increased risk for angina in New Mexico,^40^ and increased CVD mortality in France.^41^ In NHANES 2007-2008 (n=1,857), urinary uranium levels were not associated with self-reported congestive heart failure, CHD, angina, heart attack, or stroke.^42^ In the Strong Heart Family Study, a cohort of American Indians in the Southwest and Great Plains, urinary uranium levels were associated with hypertension, a major CVD risk factor.^43^ In MESA, urinary uranium levels were highest in participants from Los Angeles, CA. These findings indicate that uranium may be a significant contributor to CVD, specifically for those exposed to higher levels of uranium in the Midwest and Southwest regions. Additional research is needed to further evaluate the role of uranium in atherosclerosis and CVD development.

### Essential Metals

#### Cobalt, Co

Cobalt is an essential metal with strong ligand binding properties, low mobility, and integral as the central ion in the coenzyme vitamin B12 necessary for protein synthesis and homocysteine methylation.^44^ Cobalt is used in glass, inks, paints, and the heavy metal industry.^45^ Cobalt has been linked to non-ischemic cardiomyopathy and has experimentally shown both beneficial and deleterious effects to the cardiovascular system.^45^ Cobalt interferes with calcium binding and transport, interrupts ATP generation and production, and produces reactive oxygen species.^45^ In our study, we found that urinary cobalt levels were significantly associated with SWCS when comparing the two highest urinary cobalt quartiles to the lowest. In the Hortega study, a population-based cohort study from Spain, urinary cobalt levels were not associated with CVD risk^4^ or oxidative stress biomarkers^46^ at low levels. Our findings suggest that higher urinary cobalt levels may contribute to CAC, warranting further investigation.

#### Copper, Cu

Copper is an essential element necessary as a catalytic or structural cofactor, necessary for the regulation of oxidative stress and has been linked to CVD, particularly coronary heart disease, both when levels are deficient and in excess.^47^ Copper is widely used in agriculture as algaecides, herbicides, pesticides, wood preservation, water treatment, wiring, plumbing, and cookware.^48^ Several studies have shown PM_2.5_ composed of copper is significantly associated with increased risk of CVD and CHD,^49^ and cardiovascular mortality.^50^ A recent meta-analysis of 35 studies found that the pooled relative risk comparing the highest to lowest copper exposure tertiles was 1.81 (95% CI: 1.05, 3.11) for incident CVD and 2.22 (95%CI: 1.31, 3.74) for incident coronary heart disease.^23^ In the Hortega study, higher urinary copper levels were associated with higher CVD risk,^4^ but not with oxidative stress biomarkers.^46^ The association of urinary copper levels with SWCS at baseline and ten years of follow-up in MESA was attenuated after adjustment for CVD risk factors, with the majority of attenuation related to diabetes status. Deficient Cu levels cause increased susceptibility to LDL and HDL oxidation, a primary mechanism in the development of atherosclerosis.^51^ Excess copper can induce oxidative stress and produce reactive oxygen species, and the formation of a copper-homocysteine complex that can contribute to endothelial dysfunction and vascular injury.^23^ Although most previous studies have linked blood copper to CVD, our study shows that urinary copper levels are also linked to higher levels of CAC.

#### Zinc, Zn

Zinc is an essential element best known for its key roles in the regulation of oxidative stress, is also required for superoxide dismutase and in pancreatic islet physiology, as insulin is a hexamer made up of two zinc ions and one calcium ion. The association of urinary zinc levels with SWCS at baseline and ten years of follow-up were attenuated when adjusting for cardiovascular risk factors, including diabetes. Because zinc is an essential metal necessary for catalytic, structural, and regulatory metabolism,^52^ altered zinc homeostasis by changes in cellular zinc concentrations is an important marker of a disease state. Zinc deficiency in serum and increased urinary zinc levels have been proposed as indicators of the development of CVD and diabetes.^53^ Urinary zinc levels have been associated with oxidative stress markers^46^ and CVD incidence in the Hortega Study.^46^ Likewise, changes in cellular and free zinc ion concentrations can enhance oxidative stress.^52^ Our findings support previous evidence that urinary zinc levels are associated with CVD through increasing CAC. This may be due to higher urinary zinc levels among individuals with diabetes, a primary risk factor for CVD.

#### Clinical and Public Health implications

Our findings suggest that metals, both essential and non-essential, are related to the development of CVD at least in part through increased arterial calcification. Growing evidence from clinical trials supports that metal chelation can be beneficial for improving CVD outcomes in populations with cardiovascular disease, which could be explained by the role of chelating agents reducing non-essential metal accumulation in the body and by improving homeostasis of essential metals.^54, 55^ Given the importance of metal exposure with CVD, as supported in this study, further investigation in other large, longitudinal studies with data on CAC is necessary to further characterize this association across multiple populations, in particular to evaluate potential gene-environment interactions, characterize associations for subgroups of the population, and inform relevant interventions. These findings also provide additional support for public health actions from governments and public health agencies to lower acceptable limits of metals in air, water, and soil and improve enforcement of metal pollution reduction, particularly in communities experiencing disproportionate metal exposures.^2^ Public health interventions to reduce metal exposure may contribute to reducing CVD mortality, the leading cause of death across the globe, as supported by previous studies on the impact of lead reductions in reductions of CVD incident rates in the United States.^24^

#### Strengths and Limitations

This is a large, longitudinal study of metal exposure and subclinical CVD. Our study presents new evidence of the link between urinary biomarkers of cadmium and less studied tungsten, uranium, cobalt, copper, and zinc. Few studies of metals have assessed CAC, and most are cross-sectional. We assessed CAC prospectively to estimate changes in calcification using repeated measures of CAC, which allows to assess the association with calcification over time. To address the limitations of the dichotomized CAC-AS, we used SWCS, a more sensitive and continuous marker of calcification to maximize the available data.^20^ Our study has several limitations. Although urinary metal levels were measured in 10% of participants at Exams 1 and 5, we used urinary metal levels measured at baseline to increase power in our analysis and because levels across both exams supported that a single metal measure reflects long-term exposure and internal dose. In an exploratory analysis of the participants with time varying exposure measured at Exams 1 and 5, results were largely consistent. Residual and unknown confounding are possible, although we employed multiple sensitivity analyses using measures of ambient pollution and urinary cotinine, as well as by accounting for urine dilution using specific gravity as an alternative approach.

## Conclusions

In this prospective study of sublinical CVD across diverse urban US communities, we found that non-essential metals cadmium, tungsten, uranium, and essential metals cobalt, copper and zinc, as measured in urine, were associated with levels of CAC at baseline and over a ten year period. These findings support that atherosclerosis and resulting calcification contribute to explain the association of metals with clinical CVD outcomes. Incorporating the prevention and management of metal exposure and internal dose into clinical and public health guidelines provides novel strategies for the prevention and treatment of cardiovascular disease.

## Data Availability

Since the data is protected health information, all data is managed and released by the MESA team.

## REFERENCES

1. Martinez-Morata I, Sobel M, Tellez-Plaza M, Navas-Acien A, Howe CG, Sanchez TR. A state-of-the-science review on metal biomarkers. Current Environmental Health Reports. 2023:1–35

2. Lamas GA, Bhatnagar A, Jones MR, Mann KK, Nasir K, Tellez-Plaza M, et al. Contaminant metals as cardiovascular risk factors: A scientific statement from the american heart association. Journal of the American Heart Association. 2023:e029852

3. Grau-Perez M, Caballero-Mateos MJ, Domingo-Relloso A, Navas-Acien A, Gomez-Ariza JL, Garcia-Barrera T, et al. Toxic metals and subclinical atherosclerosis in carotid, femoral, and coronary vascular territories: The aragon workers health study. Arterioscler Thromb Vasc Biol. 2022;42:87–99

4. Domingo-Relloso A, Grau-Perez M, Briongos-Figuero L, Gomez-Ariza JL, Garcia-Barrera T, Dueñas-Laita A, et al. The association of urine metals and metal mixtures with cardiovascular incidence in an adult population from spain: The hortega follow-up study. International Journal of Epidemiology. 2019;48:1839–1849

5. Detrano R, Guerci AD, Carr JJ, Bild DE, Burke G, Folsom AR, et al. Coronary calcium as a predictor of coronary events in four racial or ethnic groups. New England Journal of Medicine. 2008;358:1336–1345

6. Kaufman JD, Adar SD, Barr RG, Budoff M, Burke GL, Curl CL, et al. Association between air pollution and coronary artery calcification within six metropolitan areas in the USA (the multi-ethnic study of atherosclerosis and air pollution): A longitudinal cohort study. Lancet (London, England). 2016;388:696–704

7. Wang M, Beelen R, Stafoggia M, Raaschou-Nielsen O, Andersen ZJ, Hoffmann B, et al. Long-term exposure to elemental constituents of particulate matter and cardiovascular mortality in 19 european cohorts: Results from the escape and transphorm projects. Environ Int. 2014;66:97–106

8. Nigra AE, Ruiz-Hernandez A, Redon J, Navas-Acien A, Tellez-Plaza M. Environmental metals and cardiovascular disease in adults: A systematic review beyond lead and cadmium. Curr Environ Health Rep. 2016;3:416–433

9. Jomova K, Valko M. Advances in metal-induced oxidative stress and human disease. Toxicology. 2011;283:65–87

10. Valko M, Morris H, Cronin MT. Metals, toxicity and oxidative stress. Curr Med Chem. 2005;12:1161–1208

11. Iavicoli I, Fontana L, Bergamaschi A. The effects of metals as endocrine disruptors. J Toxicol Environ Health B Crit Rev. 2009;12:206–223

12. Prozialeck WC, Edwards JR, Nebert DW, Woods JM, Barchowsky A, Atchison WD. The vascular system as a target of metal toxicity. Toxicol Sci. 2008;102:207–218

13. Liang CJ, Budoff MJ, Kaufman JD, Kronmal RA, Brown ER. An alternative method for quantifying coronary artery calcification: The multi-ethnic study of atherosclerosis (mesa). BMC Medical Imaging. 2012;12:14

14. Bild DE, Bluemke DA, Burke GL, Detrano R, Diez Roux AV, Folsom AR, et al. Multi-ethnic study of atherosclerosis: Objectives and design. Am J Epidemiol. 2002;156:871–881

15. Schilling K GR, Balac O, Gálvez-Fernández M, Slavkovich V, Goldsmith J, Jones MR, Sanchez TR, Navas-Acien A;. Method validation for (ultra)-trace element concentrations in urine for small sample volumes in large epidemiological studies: Application to the population-based epidemiological multi-ethnic study of atherosclerosis (mesa). 2023;Under Review

16. Nixon D, Eckdahl S, Butz J, Burrit M, Neubauer K, Schneider C. Determination of arsenic, lead, cadmium, and mercury in whole blood and urine using dynamic reaction cell icp-ms. Application Notes, Perkin-Elmer SCIEX Instruments.

17. Jaffe M. Ueber den niederschlag, welchen pikrinsäure in normalem harn erzeugt und über eine neue reaction des kreatinins. 1886;10:391–400

18. Carr JJ, Nelson JC, Wong ND, McNitt-Gray M, Arad Y, Jacobs DR, Jr., et al. Calcified coronary artery plaque measurement with cardiac ct in population-based studies: Standardized protocol of multi-ethnic study of atherosclerosis (mesa) and coronary artery risk development in young adults (cardia) study. Radiology. 2005;234:35–43

19. Nasir K, Cainzos-Achirica M. Role of coronary artery calcium score in the primary prevention of cardiovascular disease. BMJ. 2021;373:n776

20. Shea S, Navas-Acien A, Shimbo D, Brown ER, Budoff M, Bancks MP, et al. Spatially weighted coronary artery calcium score and coronary heart disease events in the multi-ethnic study of atherosclerosis. Circulation: Cardiovascular Imaging. 2021;14:e011981

21. Inker LA, Eneanya ND, Coresh J, Tighiouart H, Wang D, Sang Y, et al. New creatinine- and cystatin c–based equations to estimate gfr without race. New England Journal of Medicine. 2021;385:1737–1749

22. Kirwa K, Szpiro AA, Sheppard L, Sampson PD, Wang M, Keller JP, et al. Fine-scale air pollution models for epidemiologic research: Insights from approaches developed in the multi-ethnic study of atherosclerosis and air pollution (mesa air). Curr Environ Health Rep. 2021;8:113–126

23. Chowdhury R, Ramond A, O’Keeffe LM, Shahzad S, Kunutsor SK, Muka T, et al. Environmental toxic metal contaminants and risk of cardiovascular disease: Systematic review and meta-analysis. BMJ. 2018;362:k3310

24. Ruiz-Hernandez A, Navas-Acien A, Pastor-Barriuso R, Crainiceanu CM, Redon J, Guallar E, et al. Declining exposures to lead and cadmium contribute to explaining the reduction of cardiovascular mortality in the us population, 1988–2004. International journal of epidemiology. 2017;46:1903–1912

25. Tellez-Plaza M, Jones MR, Dominguez-Lucas A, Guallar E, Navas-Acien A. Cadmium exposure and clinical cardiovascular disease: A systematic review. Curr Atheroscler Rep. 2013;15:356

26. Majumder S, Muley A, Kolluru GK, Saurabh S, Tamilarasan K, Chandrasekhar S, et al. Cadmium reduces nitric oxide production by impairing phosphorylation of endothelial nitric oxide synthase. Biochemistry and cell biology. 2008;86:1–10

27. Biagioli M, Pifferi S, Ragghianti M, Bucci S, Rizzuto R, Pinton P. Endoplasmic reticulum stress and alteration in calcium homeostasis are involved in cadmium-induced apoptosis. Cell calcium. 2008;43:184–195

28. Lin H-C, Hao W-M, Chu P-H. Cadmium and cardiovascular disease: An overview of pathophysiology, epidemiology, therapy, and predictive value. Revista Portuguesa de Cardiologia. 2021;40:611–617

29. Ruiz-Hernandez A, Kuo CC, Rentero-Garrido P, Tang WY, Redon J, Ordovas JM, et al. Environmental chemicals and DNA methylation in adults: A systematic review of the epidemiologic evidence. Clin Epigenetics. 2015;7:55

30. Barregard L, Sallsten G, Harari F, Andersson EM, Forsgard N, Hjelmgren O, et al. Cadmium exposure and coronary artery atherosclerosis: A cross-sectional population-based study of swedish middle-aged adults. Environ Health Perspect. 2021;129:67007

31. Cullen JT, Maldonado MT. Biogeochemistry of cadmium and its release to the environment. Met Ions Life Sci. 2013;11:31–62

32. EPA (U.S. Environmental Protection Agency). Proceedings of the technical workshops for the hydraulic fracturing study: Chemical & analytical methods. 2011

33. Pang Y, Jones MR, Tellez-Plaza M, Guallar E, Vaidya D, Post WS, et al. Association of geography and ambient air pollution with urine metal concentrations in six us cities: The multi-ethnic study of atherosclerosis. Int J Environ Res Public Health. 2016;13

34. Tyrrell J, Galloway TS, Abo-Zaid G, Melzer D, Depledge MH, Osborne NJ. High urinary tungsten concentration is associated with stroke in the national health and nutrition examination survey 1999–2010. PloS one. 2013;8:e77546

35. Agarwal S, Zaman T, Murat Tuzcu E, Kapadia SR. Heavy metals and cardiovascular disease: Results from the national health and nutrition examination survey (nhanes) 1999-2006. Angiology. 2011;62:422–429

36. Guo X, Li N, Wang H, Su W, Song Q, Liang Q, et al. Combined exposure to multiple metals on cardiovascular disease in nhanes under five statistical models. Environmental Research. 2022;215:114435

37. Nigra AE, Howard BV, Umans JG, Best L, Francesconi KA, Goessler W, et al. Urinary tungsten and incident cardiovascular disease in the strong heart study: An interaction with urinary molybdenum. Environmental Research. 2018;166:444–451

38. Schnug E, Lottermoser BG. Fertilizer-derived uranium and its threat to human health. Environmental Science & Technology. 2013;47:2433–2434

39. Anderson JL, Bertke SJ, Yiin J, Kelly-Reif K, Daniels RD. Ischaemic heart and cerebrovascular disease mortality in uranium enrichment workers. Occup Environ Med. 2021;78:105–111

40. Al Rashida VJM, Wang X, Myers OB, Boyce TW, Kocher E, Moreno M, et al. Greater odds for angina in uranium miners than nonuranium miners in new mexico. J Occup Environ Med. 2019;61:1–7

41. Drubay D, Ca, xeb, r-Lorho S, Laroche P, Laurier D, et al. Mortality from circulatory system diseases among french uranium miners: A nested case-control study. Radiat Res. 2015;183:550–562

42. Mendy A, Gasana J, Vieira ER. Urinary heavy metals and associated medical conditions in the us adult population. Int J Environ Health Res. 2012;22:105–118

43. Gold AON-A, Ana. Assessment of differential uranium exposure and its association with hypertension and elevated blood pressure in american indian communities in the strong heart family study. 2023;2023

44. Jomova K, Makova M, Alomar SY, Alwasel SH, Nepovimova E, Kuca K, et al. Essential metals in health and disease. Chemico-Biological Interactions. 2022;367:110173

45. Packer M. Cobalt cardiomyopathy. Circulation: Heart Failure. 2016;9:e003604

46. Domingo-Relloso A, Grau-Perez M, Galan-Chilet I, Garrido-Martinez MJ, Tormos C, Navas-Acien A, et al. Urinary metals and metal mixtures and oxidative stress biomarkers in an adult population from spain: The hortega study. Environment International. 2019;123:171–180

47. Klevay LM. Cardiovascular disease from copper deficiency--a history. J Nutr. 2000;130:489s–492s

48. Agency for Toxic Substances and Disease Registry (ATSDR). Toxicological profile for copper. 2022

49. Badaloni C, Cesaroni G, Cerza F, Davoli M, Brunekreef B, Forastiere F. Effects of long-term exposure to particulate matter and metal components on mortality in the rome longitudinal study. Environ Int. 2017;109:146–154

50. Lavigne A, Freni Sterrantino A, Liverani S, Blangiardo M, de Hoogh K, Molitor J, et al. Associations between metal constituents of ambient particulate matter and mortality in england: An ecological study. BMJ Open. 2019;9:e030140

51. James JD, Dennis M, James HOK. Copper deficiency may be a leading cause of ischaemic heart disease. Open Heart. 2018;5:e000784

52. Maret W. Zinc in pancreatic islet biology, insulin sensitivity, and diabetes. Prev Nutr Food Sci. 2017;22:1–8

53. Galvez-Fernandez M, Powers M, Grau-Perez M, Domingo-Relloso A, Lolacono N, Goessler W, et al. Urinary zinc and incident type 2 diabetes: Prospective evidence from the strong heart study. Diabetes Care. 2022;45:2561–2569

54. Ravalli F, Vela Parada X, Ujueta F, Pinotti R, Anstrom KJ, Lamas GA, et al. Chelation therapy in patients with cardiovascular disease: A systematic review. Journal of the American Heart Association. 2022;11:e024648

55. Lamas GA, Goertz C, Boineau R, Mark DB, Rozema T, Nahin RL, et al. Effect of disodium edta chelation regimen on cardiovascular events in patients with previous myocardial infarction: The tact randomized trial. Jama. 2013;309:1241–1250

